# Early versus Later Anticoagulation in Post-Stroke patients with Atrial Fibrillation: A systematic review and Meta-analysis

**DOI:** 10.1101/2024.04.15.24305864

**Authors:** Carlos Zavaleta-Corvera, Gino Vasquez-Paredes, Joaquín Sarmiento-Falen, Olmedo Riveros-Hernandez, José Caballero-Alvarado

## Abstract

**Background:** Atrial fibrillation (AF) is the most common cardiac arrhythmia and significantly increases the risk of ischemic stroke, one of its most serious complications. The intricate relationship between AF and stroke necessitates careful consideration in the timing of anticoagulant therapy initiation to prevent stroke recurrence, hemorrhagic transformation, and other complications.

**Aim:** This study aims to evaluate the efficacy and safety of early versus late initiation of anticoagulant treatment in patients with AF after an ischemic stroke, addressing the critical gap in optimal timing to balance the risk of recurrent stroke against bleeding risks.

**Methods:** A systematic review and meta-analysis was conducted following the PRISMA-2020 guidelines, with searches performed across multiple databases including PubMed, Scopus, Web of Science, EMBASE, and Cochrane. Randomized controlled trials comparing early versus later initiation of anticoagulation in AF patients post-stroke were included. Data synthesis was performed using a random-effects model, considering recurrent ischemic stroke, symptomatic intracerebral hemorrhage, and mortality as primary outcomes.

**Results:** From 1444 identified records, three studies and 2989 patients were included, comprising a diverse population in terms of geography and demographics. Meta-analysis revealed that early anticoagulation initiation might be associated with a reduced risk of recurrent ischemic stroke (RR 0.72, 95% CI [0.51-1.00])(p=0.05), although statistical significance was borderline. No significant differences were found in symptomatic intracerebral hemorrhage or mortality rates between early and late initiation. Early intervention was, however, associated with a reduction in other adverse outcomes (RR 0.72, 95% CI [0.54-0.96])(p=0.03).

**Conclusions:** Early initiation of anticoagulation therapy in AF patients post-ischemic stroke may offer a protective effect against recurrent ischemic events with no significant increase in bleeding or mortality risks. This suggests that early anticoagulation can be safely considered, particularly in light of reducing other adverse outcomes.

## INTRODUCTION

Atrial fibrillation (AF) is the most common cardiac arrhythmia, substantially increases the risk of ischemic stroke, representing one of its most serious complications (1,2). Both conditions are closely related and have been suggested to be mutually causal (3). Feared complications of AF-induced ischemic stroke include stroke recurrence, haemorrhagic transformation, cognitive impairment, functional decline, and cardiovascular complications (4-8). Consequently, the timing of initiating anticoagulant therapy in patients diagnosed with AF who have suffered an ischemic stroke emerges as a critical clinical question to prevent these outcomes (9).

Fundamental to their effectiveness is the ability of anticoagulants to alter key factors within the coagulation cascade, significantly reducing the risk of clot formation that could precipitate ischemic events (10). The therapeutic landscape is populated by a wide range of anticoagulants, including well-established vitamin K antagonists such as warfarin and more recently developed direct oral anticoagulants (DOACs) such as dabigatran, rivaroxaban, and apixaban (11). The advent of DOACs marks a paradigm shift and offers clear advantages over traditional therapies, particularly through an improved safety profile (12, 13). This improvement not only simplifies patient management but also reduces the need for frequent monitoring. Together, these benefits contribute to promoting greater adherence to treatment protocols (14). Selection of an appropriate anticoagulant is nuanced and requires judicious evaluation of several patient-specific factors, including, but not limited to, renal function, concomitant drug interactions, and the individual’s bleeding risk profile (15,16).

The unsolved clinical challenge (as reported by 95% of stroke doctors in the UK) lies in deciding when to start anticoagulation, given the high risk of recurrence (0.5% to 1.3% per day, which increases with age and extensive infarctions) (17). In addition to considering the possible complications of anticoagulation (eg, intracerebral hemorrhage, 9%) (18). Some current evidence recommends delaying anticoagulation in patients with large infarcts and suggests performing a brain CT scan before initiating anticoagulation (19-21).

However, the timing of initiating anticoagulation in patients with stroke and atrial fibrillation (AF) requires a delicate balance between preventing recurrent stroke and minimizing bleeding risks. Early initiation of anticoagulant treatment, usually within the first 14 days after stroke, has been associated with a reduced risk of recurrent stroke (22). Nevertheless, this strategy may pose a high risk of hemorrhagic transformation of ischemic stroke, particularly in the first few days after stroke (23,24).

On the other hand, delaying anticoagulant therapy beyond this time frame may facilitate stabilization of the infarct area and restoration of the blood-brain barrier, potentially mitigating the risk of haemorrhagic complications (25). However, each day without anticoagulant treatment widens the window of vulnerability to recurrent strokes, underscoring the critical importance of timely intervention in stroke prevention in patients with AF (26). The American Heart Association/American Stroke Association guidelines emphasize the need for personalized approaches based on individual patient characteristics in the treatment of acute ischemic stroke (AIS) (27).

The aim of the present study will be to evaluate the efficacy and safety of early versus late initiation of anticoagulant treatment in patients with atrial fibrillation (AF) after ischemic stroke.

## METHODS

### Study Design

We conducted a Systematic Review with meta-analysis design. The PRISMA-2020 (Preferred Reporting Items for Systematic Reviews and Meta-Analysis) recommendations served as the review process parameters. PROSPERO codification (Prospective International Registry of Systematic Reviews): CRD42024528280.

### Search Strategy

There was a database search using PubMed, Scopus, Web of Science, EMBASE and Cochrane. MeSH phrases about a combination of terms related to “Atrial Fibrillation,” “Stroke,” “Anticoagulation,” and Boolean operators (AND, OR) were used to combine search terms until 1st March 2024. Results were not limited by language or date of publication. In addition, all reference lists of systematic reviews and included review articles were hand-searched for other potentially suitable articles. The search strategy was peer-reviewed by an expert in systematic review methodologies using the Peer Review of Electronic Search Strategies (PRESS) guideline.

### Eligibility Criteria

Only original articles that meet the following criteria were included in this study: Randomized controlled trials (RCTs). This selection is aimed at adults diagnosed with atrial fibrillation who had subsequently experienced an ischemic stroke. The interventions in question compared the timing of anticoagulation therapy initiation standardized in the following definitions: Early initiation, defined as within initiation of anticoagulation therapy within 48 hours of stroke onset or within ≤4 days from stroke onset, aimed at preventing further blood clot formation and reducing the risk of recurrent strokes during the acute phase of stroke management. Later Anticoagulation, defined as Initiation of anticoagulation therapy beyond the acute phase of stroke management, typically starting 5–10 days after stroke onset or on specific days post-stroke, such as day 3 or 4 after a minor stroke, day 6 or 7 after a moderate stroke, or day 12, 13, or 14 after a major stroke.

We excluded non-RCTs and observational studies such as cohort studies, case-control studies, cross-sectional studies, descriptive studies, and quasi-randomized trials. Animal studies and non-original research articles (including conference abstracts, reports, case series, systematic reviews, narratives, and letters to the editor) will also be omitted from our analysis. Importantly, the date of publication will not serve as a basis for exclusion, allowing for a comprehensive and historical perspective on the available evidence.

### Outcomes

#### Primary Outcomes

The first primary outcome is recurrent ischemic stroke, which is identified as a new episode of ischemic stroke occurring after the initial stroke event. Confirmation of a recurrent ischemic stroke is obtained through neuroimaging or clinical assessment, ensuring that only definitive cases are considered. The second primary outcome is symptomatic intracerebral hemorrhage, characterized by bleeding within the brain parenchyma that manifests with clinical symptoms. Like the identification of recurrent ischemic stroke, symptomatic intracerebral hemorrhage is confirmed through neuroimaging, allowing for accurate and reliable diagnosis.

#### Secondary Outcomes

Mortality for all the causes, defined as death from any cause during the study follow-up period, will be tracked to assess the ultimate impact of therapy timing on patient survival. Others to be considered were composite outcomes, where we included all the reports in the articles analysed.

### Study Selection and Data Collection Process

Systematic searches were conducted in the following electronic databases: PubMed, Scopus, Web of Science, EMBASE and Cochrane. The search strategy was adapted for each database to capture all relevant studies (Appendix 1). Searches were complemented by checking the reference lists of included studies and relevant reviews, as well as consulting clinical trial registries. Two reviewers independently screened titles and abstracts for eligibility. Full texts of potentially eligible studies retrieved and independently assessed for inclusion by the same reviewers. Disagreements were resolved through discussion or consultation with a third reviewer. A standardized data extraction form was used to collect information from included studies on study characteristics, participant demographics, interventions, outcomes, and risk of bias. Data extraction was conducted independently by two reviewers, with discrepancies resolved by consensus or a third reviewer.

### Risk of Bias Assessment

The RoB 2.0 tool (Risk of Bias 2.0) was used to evaluate the risk of bias. Any differences of opinion will be settled by conversation or advice from a third author. The results had three categories: low, moderate concerns, and high risk of bias.

Thus, several domains of bias were assessed, such as randomization, concealed allocation, blinding of participants and staff, incomplete data management, and selective reporting of results. Each domain was assessed individually to determine whether there is a high risk of bias, some problems, or a low risk of bias. Disagreements in the assessment of each domain were addressed by discussion among the assessors or by consultation with a third author, which ensured objectivity and accuracy in assessing the risk of bias in the study.

### Data Synthesis

Software and Models: Meta-analyses were conducted using the latest version of R software. Given the expected variability between studies regarding patient characteristics, interventions, and outcomes, a random-effects model was employed for all analyses to account for between-study heterogeneity. The inverse variance method served as the primary analytical technique for combining study results.

Outcome Measures: The effects of early versus later initiation of anticoagulation on dichotomous outcomes, such as recurrent ischemic stroke and symptomatic intracerebral hemorrhage, were described using risk ratios (RRs) with 95% confidence intervals (CIs). For continuous outcomes, such as functional outcomes and quality of life scores, mean differences (MD) or standardized mean differences (SMD) with 95% CIs were calculated, depending on the scales used across studies.

Handling Heterogeneity and Variance: To quantify heterogeneity among included studies, the I² statistic was used, with I² values of 25%, 50%, and 75% indicating low, moderate, and high heterogeneity, respectively. The Paule-Mandel estimator was used to calculate the between-study variance (tau²) in the random-effects model. For studies reporting zero events in one or both arms, a continuity correction was applied to allow for inclusion in the meta-analysis.

Sensitivity Analysis and Publication Bias: Sensitivity analyses were conducted to explore the robustness of the findings by excluding studies with high risk of bias or using alternative statistical models, such as fixed-effects models for comparison. To assess the potential for publication bias, funnel plots were visually inspected, and Egger’s test was conducted if the number of included studies allowed (typically ten or more studies).

Subgroup Analyses: Where data permitted, subgroup analyses were performed based on key study characteristics that may influence the outcomes, such as the type of anticoagulant used (e.g., direct oral anticoagulants vs. warfarin), patient age, stroke severity, and timing of anticoagulation initiation (early defined as within 48 hours to 7 days post-stroke, and later defined as beyond 7 days post-stroke).

Adjustments for Multiple Comparisons: Given the multiple outcomes assessed in this meta-analysis, results were interpreted cautiously, considering the potential for type I error due to multiple comparisons.

### GRADE Assessment

The GRADE system was employed to evaluate the degree of certainty in the evidence and develop suggestions. During this process, a number of domains, such as publication bias, indirectness, inconsistency, risk of bias, and imprecision, will be examined. Furthermore, the GRADEpro GDT tool was utilized to generate Summary of Findings (SoF) tables.

### Ethics and Dissemination

As this is a systematic review, ethical approval is not required.

## RESULTS

A total of 1444 relevant studies were initially identified through five major databases: PubMed (n=279), Embase (n=131), Web of Science (n=78), Scopus (n=923) and Cochrane (n=33). After, we eliminated 233 duplicate records, 527 ineligible due to automation tools, 569 because they were not clinical trials or did not focus on the specific topic and 112 not recovered or due to lack of complete data or the absence of full text available. Thus, three studies were included in the analysis.

After full-text screening, three studies with a sample size of 2989 patients were included in the systematic review. The Fischer 2023 (ELAN) study (28), carried out in 15 countries, analyzed 2013 participants with a median age of 77 years (range 70 to 84 years), standing out for its wide geographical coverage and focus on elderly people, with a majority of 55.4% male. Oldgren 2022 (TIMING) (29), focused on Sweden and based on registries, included 888 patients, showing a mean age of 78.3 years (standard deviation 9.9 years) and a slight male preponderance of 53.8%. Labovitz 2021 (AREST) (30), a controlled trial in the US, had 88 participants, focusing on a more balanced gender distribution with 44.3% men and a mean age of 73.5 years (standard deviation 12.7 years). These studies reflect varied methodologies and geographic contexts, underlining the importance of post-stroke anticoagulation strategies in older adults, demonstrating the need to adapt interventions to the demographic characteristics of the population internationally(Table 1).

**Table 1.**
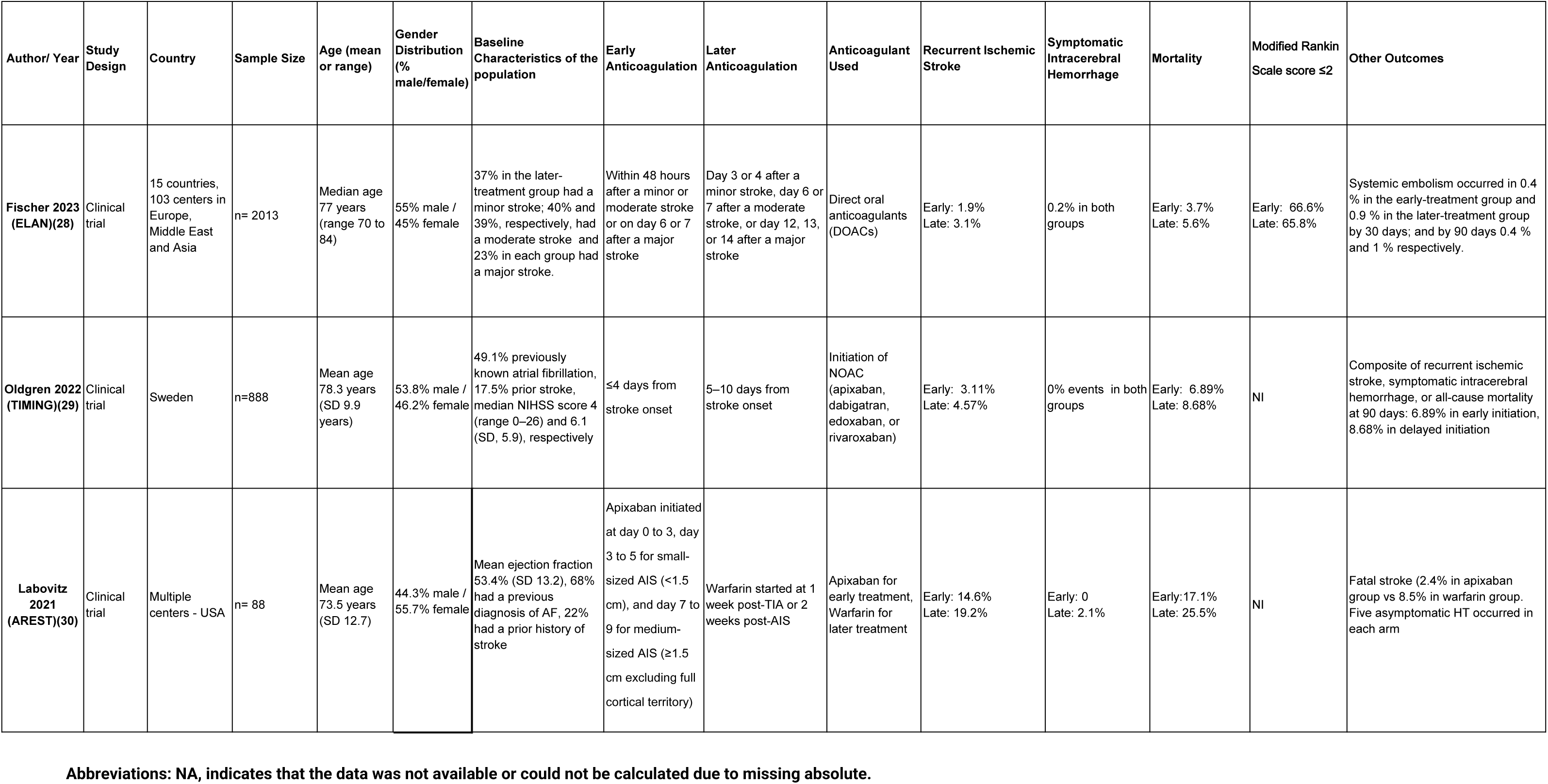
Summarize of the articles included.

### Recurrent Ischemic Stroke

We considered for the data extraction the maximum time of evaluation of this outcome. The fixed-effects meta-analysis yields a combined relative risk (RR) across all studies is 0.72 (95% CI [0.51-1.00]). There is minimal heterogeneity among the included studies, with an I-squared value of 0%, and the Chi-square statistic does not suggest significant variability (Chi-square = 0.49, degrees of freedom = 2, p = 0.78). The overall effect test yields a z-value of 1.96 (p = 0.05), indicating that the overall effect size is at the threshold of conventional levels of statistical significance (Figure 1).

**Figure 1.**
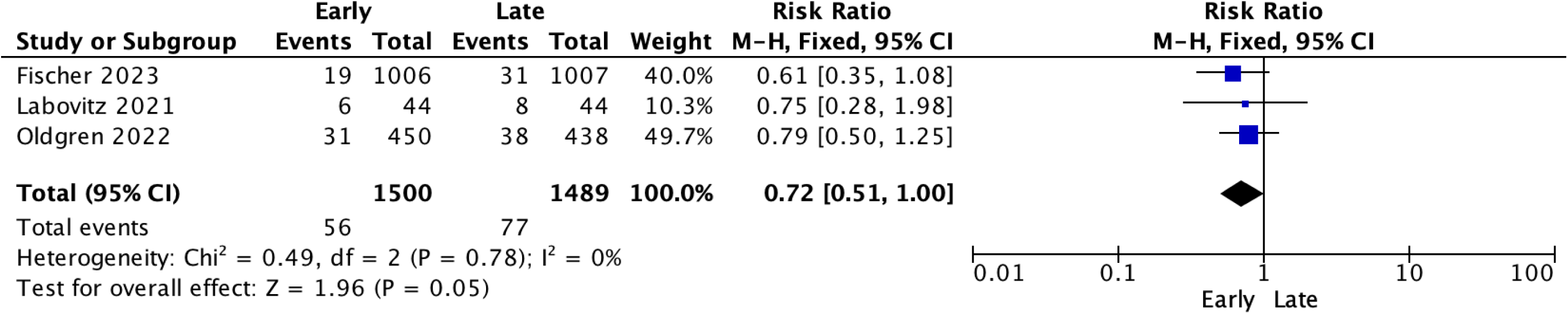
Recurrent Ischemic Stroke Forest Plot.

### Symptomatic Intracerebral Hemorrhage

The fixed-effects meta-analysis yields a combined RR of 0.71 (95% CI [0.14-3.60]). The analysis shows minimal heterogeneity, with an I-squared (I²) value of 0% and a Chi-squared statistic of 0.34 (p = 0.56), indicating consistency across study results. Additionally, the overall effect as indicated by the z-value is 0.41(p=0.68), which does not reach the conventional level of statistical significance. This suggests that there is no compelling statistical evidence of a difference in the risk of symptomatic intracerebral hemorrhage between early and late anticoagulation based on the current data (Figure 2).

**Figure 2.**
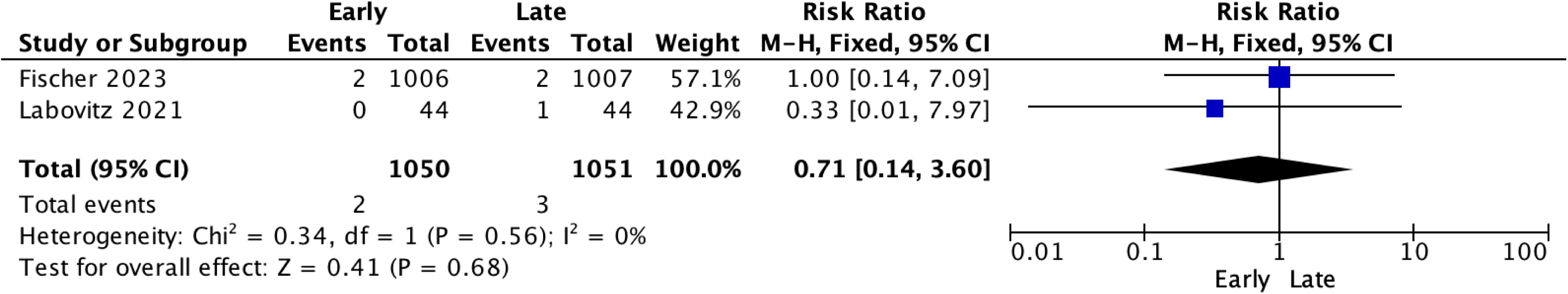
Symptomatic Intracerebral Hemorrhage Forest Plot.

### Mortality

The combined risk ratio in all studies is 0.81 (95% CI [0.59 to 1.12]). There appears to be insignificant heterogeneity between the studies, as indicated by an I-squared (I²) value of [0%] and a non-significant Chi-squared statistic of 0.35 (p = 0.84). The overall effect test yields a Z-score of 1.26 with a (p=0.21), suggesting that there is no statistically significant difference in mortality between early and late anticoagulation based on this combined analysis (Figure 3).

**Figure 3.**
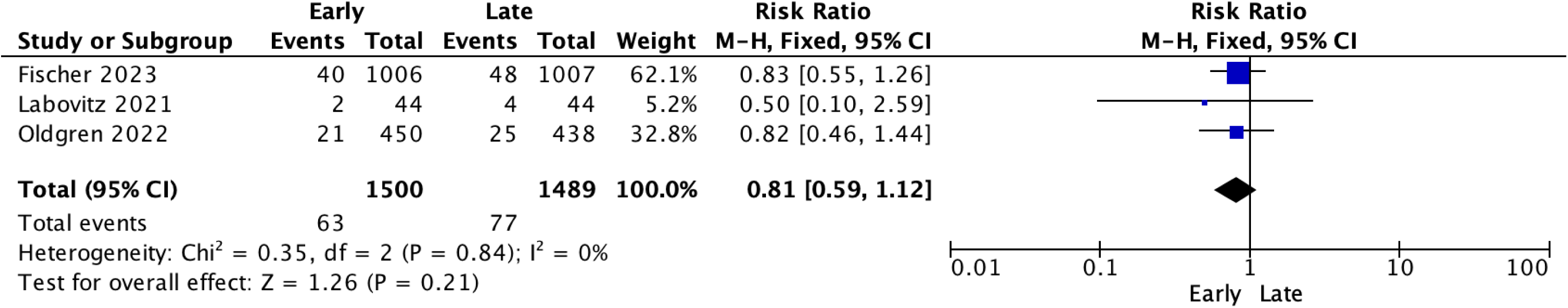
Mortality Forest Plot.

### Composite Outcomes

It shows that in the early treatment group, there were 75 events among 1500 participants, while the late treatment group had 103 events among 1489 participants. The meta-analysis yields a pooled RR of 0.72 (95% CI [0.54-0.96]). The homogeneity of the studies is reflected by an I-squared (I²) of 0% and a Chi-square statistic of 0.31 (p=0.86), indicating a lack of significant variability among study results. The test for overall effect has a Z-score of 2.24 (p=0.03) indicating a statistically significant difference in the incidence of adverse reactions between the early and late anticoagulation groups, with early treatment associated with fewer such events (Figure 4).

**Figure 4.**
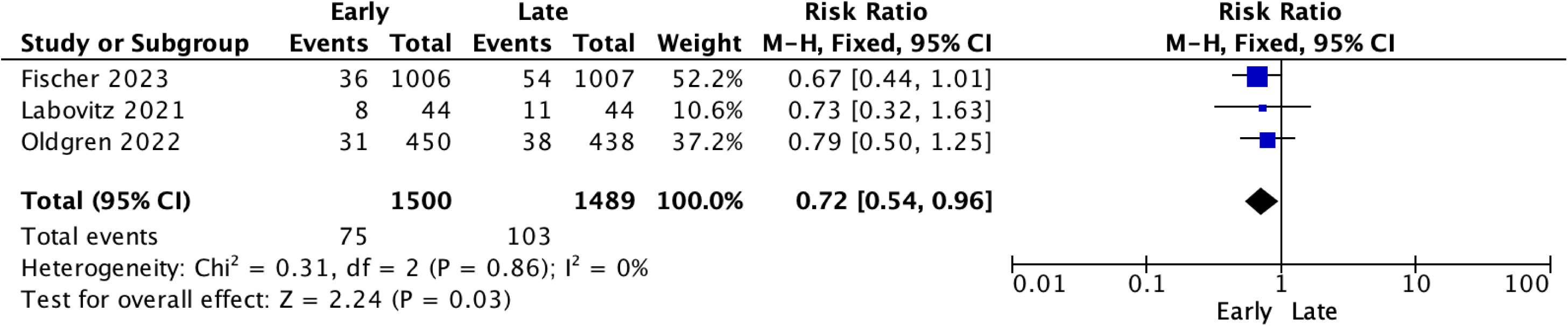
Composite Outcome - Other Adverse reactions Forest Plot.

### Bias risk analysis

The analysis of the risk of Bias was carried out through the use of the ROB2.0 tool. Thus, five domains called D1 to D5 were evaluated. Fishcer 2023 and Oldgren 2022 suggest a low risk of bias, however, Labovitz 2021 showed yellow symbols in domains D1 and D2. In assessing the distribution of overall risk levels across studies, factors such as overall bias, selection of reported outcome, outcome measurement, missing outcome data, deviations from intended interventions were assessed. and the randomization process. The graph showed most assessments at low risk (green), with some concerns (yellow) represented in smaller proportions. In particular, there are no high-risk (red) assessments in any category. This suggests that overall the body of evidence being evaluated has a low risk of bias, although there are specific areas where some studies raise concerns(Figure 5).

**Figure 5.**
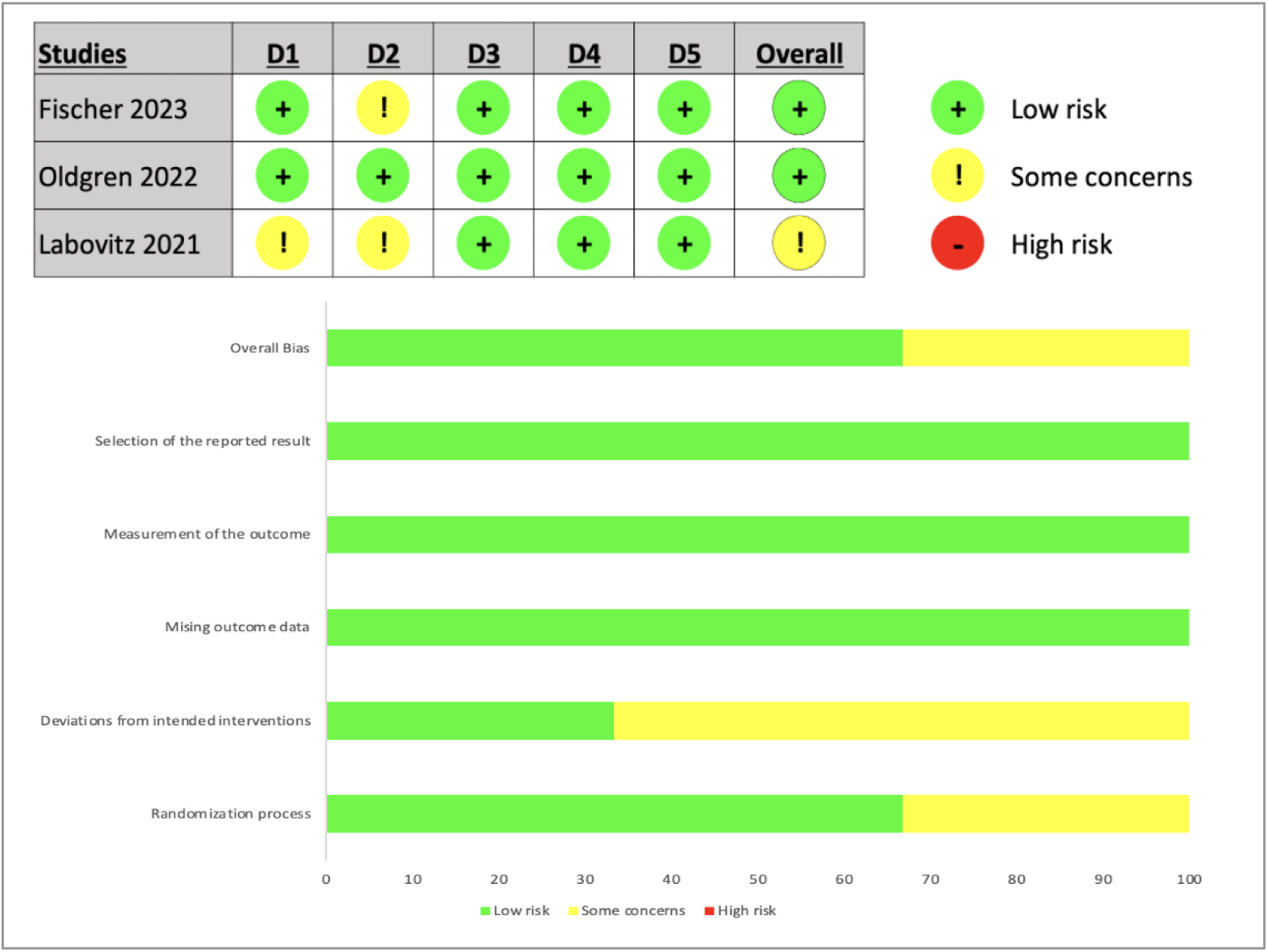
Risk of Bias Assessment with Rob2 tool.

### GRADE analysis

The GRADE certainty of evidence assessment across the AREST (2021)(30), TIMING (2022)(29), and ELAN (2023)(28) trials highlights a landscape of anticoagulation therapy timing in atrial fibrillation patients post-acute ischemic stroke. Specifically, the outcomes such as recurrent stroke/TIA and symptomatic intracerebral hemorrhage from the AREST and ELAN trials show a spectrum from low to high certainty, emphasizing the need for precise clinical judgment in the early initiation of therapy. Although these trials collectively suggest potential benefits in early anticoagulation, particularly with direct oral anticoagulants, the varying degrees of evidence underscore the complexity of clinical decision-making in this context, balancing the risks of recurrent ischemic events against those of bleeding (Table 2).

**Table 2.** Certainty of GRADE evidence.

| Outcomes | No. of participants (studies) | Follow-up | Certainty of Evidence (GRADE) | Relative Effect (95% CI) | Anticipated Absolute Effects |
| --- | --- | --- | --- | --- | --- |
| Recurrent Stroke/TIA | AREST: 88 (1 RCT), TIMING: 888 (1 RCT), ELAN: 2013 (1 RCT) | AREST: Not specified, TIMING: 90 days, ELAN: 90 days | Low (AREST) ⊕○○○, Moderate (TIMING) ⊕⊕○○, Moderate (ELAN) ⊕⊕○○ | AREST: Not provided, TIMING: Early: 3.11% vs. Delayed: 4.57%, ELAN: Early: 1.9% vs. Delayed: 3.1% | AREST: Not provided, TIMING: Risk with Delayed: 4.57%, ELAN: Risk with Delayed: 3.1% |
| Death | AREST: 88 (1 RCT), TIMING: 888 (1 RCT), ELAN: Not reported | AREST: Not specified, TIMING: 90 days, ELAN: Not specified | Low (AREST) ⊕○○○, Moderate (TIMING) ⊕⊕○○ | AREST: Not provided, TIMING: Early: 4.67% vs. Delayed: 5.71%, ELAN: Not provided | AREST: Not provided, TIMING: Risk with Delayed: 5.71%, ELAN: Not provided |
| Symptomatic Intracerebral Hemorrhage | AREST: 88 (1 RCT), TIMING: 888 (1 RCT), ELAN: 2013 (1 RCT) | AREST: Not specified, TIMING: 90 days, ELAN: 30 days | Moderate (AREST) ⊕⊕○○, High (TIMING) ⊕⊕⊕⊕, High (ELAN) ⊕⊕⊕⊕ | AREST: Not provided, TIMING: Early: 0% vs. Delayed: 0%, ELAN: Early: 0.2% vs. Delayed: 0.2% | AREST: Not provided, TIMING: Risk with both: 0%, ELAN: Risk with both: 0.2% |
| Composite Outcome | AREST: 88 (1 RCT), TIMING: 888 (1 RCT), ELAN: 2013 (1 RCT) | AREST: Not specified, TIMING: 90 days, ELAN: 30 days | Low (AREST) ⊕○○○, Moderate (TIMING) ⊕⊕○○, High (ELAN) ⊕⊕⊕⊕ | AREST: Not provided, TIMING: Early: 6.89% vs. Delayed: 8.68%, ELAN: Early: 2.9% vs. Delayed: 4.1% | AREST: Not provided, TIMING: Risk with Delayed: 8.68%, ELAN: Risk with Delayed: 4.1% |

## DISCUSSION

Regarding sociodemographic information analysed. We found a mean age of 76.26 years. Multiple evidence suggests from much time ago a relationship between age and the incidence of stroke in patients with atrial fibrillation. Sun Y, et al. in 2021 found in a cohort study that being an older patient (>=55 years) is one of the most important risk factor for stroke in atrial fibrillation: OAC therapy vs non-OAC Treatment (males HR 0.66, IC 0.49 -0.84, p=0.011 and females HR 0.69, 95% CI 0.51 – 0.87, p=0.013)(31). And other hand Tae -Hoon K, et al, 2017 found in a clinical trial that females 65-74 years had a lower risk of ischemic stroke than males (HR 0.73, 95%, CI 0.64 – 0.84), with an event rate < 1% per year(32).

Also, we found a statistically significant reduction in recurrent ischemic stroke with early anticoagulation initiation in post-stroke patients with atrial fibrillation. Tirumandyam G, et al. (2023) underscore this benefit, showing early treatment significantly reduces stroke recurrence (RR: 0.71, 95% CI: 0.53 - 0.94, p-value: 0.02), without elevating mortality or hemorrhage risks(33). Further reinforcing our findings, De Marchis et al. (2021) analysis indicates that early versus late DOAC initiation does not significantly alter the risk of recurrent AIS (aHR: 1.2, 95% CI: 0.5 - 2.9, p-value: 0.69), affirming the safety and effectiveness of initiating anticoagulation early(34).

Regarding to symptomatic intracerebral hemorrhage between early and late anticoagulation we didn’t find statistical evidence of a difference in the risk of based on the current data. Wilson D, et al. differs about our findings. They found that patients who were started early in anticoagulation developed lower numbers of have a significant haemorrhagic transformation or symptomatic intracranial bleeding (35).

Funrthermore, our alnalysis suggested that there were not statistically significant differences in mortality between early and late anticoagulation based on this combined analysis. Tirumandyam G, et al. (2023) analyzed all-cause mortality between the early and delayed anticoagulation through a systematic review and meta analysis in which didn’t find significant differences between groups (RR: 0.71, 95% CI: 0.40-1.28, p-value: 0.26)(33). Wilson D, et al. (2019) through a cohort study found no significant result between early or later anticoagulation in patients with ischemic stroke after AF (OR starting late 0.91, 95% CI [0.20-4.60])(35). In spite of all studies there were no significant results, low mortality rates were reported in all the studies reviewed(28,29,30,1,2).

As for the composite outcome of other adverse reactions, it was statistically significant, showing that the risk is significantly lower in patients in the early anticoagulation group compared to the late group. These results are comparable with the study by Tirumandyam G, et al. where they demonstrated that the risk of a composite outcome is lower in participants who received early anticoagulation compared to their counterparts. (RR: 0.69, 95% CI: 0.51-0.93, p-value: 0.01). No significant heterogeneity in study outcome was reported. Similarly in the study by Gayathri et al., (I-squared: 6%, p value: 0.34)(33).

In another multicenter observational cohort study (CROMIS-2), Wilson D, et al. suggested that early anticoagulation after ischemic stroke in patients with atrial fibrillation did not lead to a difference in the composite outcome compared with late anticoagulation. This highlights the safety of starting early anticoagulation without adding risks of adverse reactions to the composite result(35).

## CONCLUSIONS

The analysis of the studies shows that early anticoagulation could be associated with a lower risk of recurrent ischemic stroke, although statistical significance is borderline. No significant differences were observed in symptomatic intracranial cerebral hemorrhage between the intervention periods. Regarding all-cause mortality, there are no significant benefits of early intervention compared to late intervention. However, early intervention is associated with a reduction in other composite outcomes.

## Data Availability

All data referenced in this manuscript are available upon request from the corresponding author.

## Ethical aspects

All authors certify that they meet the current authorship criteria of the International Committee of Medical Journal Editors (ICMJE).

## Declaration of conflict of interest

The authors have no conflict of interest to declare.

## Funding Sources

The authors declare that this work has not received any funding from funding agencies in the public, commercial, or non-profit sectors.

## Author Contributions

Zavaleta-Corvera C, Vasquez-Paredes G, Sarmiento-Falen J, Riveros-Hernandez O: Conceptualization, Formal research analysis, Methodology, Resources, Software, Validation, Visualization, Approval of the final manuscript.

Caballero-Alvarado J: Writing, Review and editing, Approval of the final manuscript.

---

**Graphic 1.**
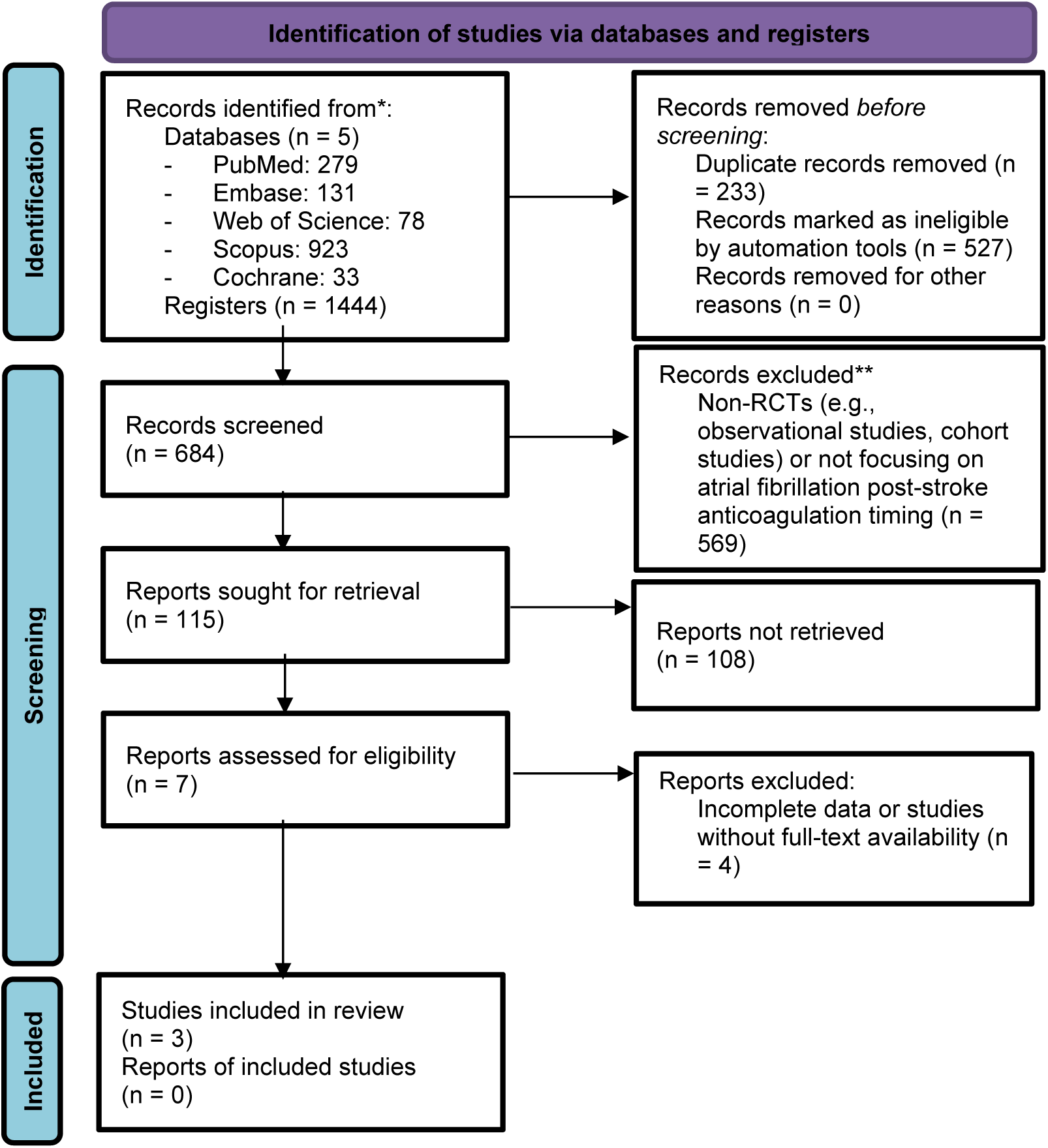
PRISMA 2020 flow diagram for new systematic reviews which included searches of databases and registers only.

## Notes

### Competing Interest Statement

The authors have declared no competing interest.

### Funding Statement

None

